# Cumulative Social Disadvantage and Health-Related Quality of Life: National Health Interview Survey 2013-2017

**DOI:** 10.1101/2022.08.20.22278956

**Authors:** Kobina Hagan, Zulqarnain Javed, Miguel Cainzos-Achirica, Adnan A. Hyder, Elias Mossialos, Tamer Yahya, Isaac Acquah, Javier Valero-Elizondo, Alan Pan, Nwabunie Nwana, Mohamad Taha, Khurram Nasir

## Abstract

**Background:** Evidence of the association between social determinants of health (SDoH) and health-related quality of life (HRQoL) is largely based on single SDoH measures, with limited evaluation of cumulative social disadvantage. We examined the association between cumulative social disadvantage and the Health and Activity Limitation Index (HALex).

**Methods:** We used data of respondents from the 2013-2017 National Health Interview Survey. A cumulative SDoH index was created by aggregating 46 SDoH from 6 domains, and respondents were grouped by quartiles (SDoH-Q1 to Q4). A higher SDoH index or quartile indicates greater disadvantage. Two outcomes were assessed: HALex score using two-part models, and a binary “poor HALex” (HALex score < 0.79 [20^th^ percentile]) using logistic regression. Regression analyses adjusted for demographics and comorbidities and were performed in the overall sample, and by age, sex, and race/ethnicity categories.

**Results:** Among 156,182 adults (mean age 46 years; 51.7% women), higher SDoH quartile groups averaged lower HALex scores and had higher proportions with poor HALex. A unit increase in SDoH index was associated with 0.01 decrease in HALex score (β = −0.01; 95% CI [−0.01, −0.01]) and 14% higher odds of poor HALex (odds ratio, OR = 1.14; 95% CI [1.14, 1.15]). Relative to SDoH-Q1, SDoH-Q4 was associated with HALex score decrease of −0.13 (95% CI [−0.13, −0.13]) and OR 8.67 (95% CI [8.08, 9.30]) for poor HALex. Hispanic persons, despite a relatively higher burden of cumulative social disadvantage, had a weaker SDoH-HALex association than their counterparts.

**Conclusion:** Higher cumulative social disadvantage was independently associated with lower HRQoL in an incremental fashion. The more favorable HRQoL profile observed in the Hispanic could be related to a resilient concept of health derived from cultural values and acceptance, and this highlights the need for population “wellness” interventions to be responsive to such phenomenon.

## Background

Patient-reported outcomes have gained traction in the evaluation of quality healthcare and monitoring of population health in recent years (1,2). These subjective outcomes directly inform providers and health policy-makers about the experiences of patients (or groups) and their value systems, both of which influence therapeutic choices, disease management, and health policy. Health-related quality of life (HRQoL), a commonly assessed patient-reported outcome measure, captures symptoms and functional limitations associated with health conditions and is known to be influenced by an individual’s psychosocial environment and their value-based preferences (3).

Likewise, the role of social determinants of health (SDoH) in perpetuating disparities in health and healthcare delivery has garnered attention as well (4–6). SDoH are observed to influence objective health outcomes such as morbidity and mortality (7–9), and patient-reported health and wellbeing (10,11). SDoH are intricately interconnected and adverse determinants often cluster in marginalized groups (12–14). Additionally, these proximal determinants can have differential impact on health which calls for a multidimensional framework in their assessment. Yet, evidence on the association between SDoH and HRQoL is largely based on single assessments of SDoH (11,15,16). Very few indices of cumulative social disadvantage capture burden across an exhaustive range of SDoH domains (17–19). A comprehensive multidimensional SDoH framework may afford a provider or health system nuanced information to better identify at-risk individuals or those likely amenable to social interventions.

Thus, in this cross-sectional study, we utilized a framework of multiple SDoH from 6 domains (20) to evaluate the Health and Activity Limitation Index (HALex) (21). HALex is a generic HRQoL measure combining self-reported health with the performance of activities of daily living and instrumental activities of daily living (21). It summarizes a person’s HRQoL into a global score ranging from 0 (dead) to 1.00 (perfect health and functioning). Our choice of HALex was informed by its unique feature to combine two different dimensions of health status, unlike other generic measures which separately assess these dimensions (22). We assessed HALex scores across levels of cumulative social disadvantage for the total population and by age, sex, and race/ethnicity. We hypothesized that higher levels of cumulative social disadvantage would be associated with poorer performance on HALex.

## Methods

### Data Source and Study Sample

The National Health Interview Survey (NHIS) is a cross-sectional household interview survey conducted annually by the National Center for Health Statistics (23). A complex multistage area probability design is used to sample the non-institutionalized civilian population of the United States. Survey items are organized into four core components – Household Composition, Family, Sample Child, and Sample Adult. The Household Composition core collects basic demographic and relationship information about all persons in the household. The Family Core, administered separately for each family in the household, collects information on sociodemographic characteristics, indicators of health status, activity limitations, injuries, health insurance coverage, and access to and use of health services. From each family, one sample child and one sample adult are randomly selected to gather further information on them. In this study, we supplemented the Sample Adult core with variables from the Family and Household Composition files.

Since the survey years 2013 to 2017 consistently featured items for our SDoH variables, we used data from these survey years to limit missingness. The data of respondents aged ≥18 years were merged and pooled over the five years. Survey weights were used to account for selection probability and non-response. NHIS data files are publicly available and deidentified, hence this study was exempt from the purview of the Houston Methodist’s Institutional Review Board.

### Study Measures

#### Independent Variable

Our cumulative SDoH index was developed using information for 46 social factors selected and organized into 6 domains as proposed by the Kaiser Family Foundation SDoH framework □ economic stability, neighborhood/physical environment, education, food security, community/social context, and healthcare system (**Figure 1**) (20). **Supplemental Table S1** contains the survey items used to obtain the SDoH information. Each item was scored as “0” if favorable and “1” if “unfavorable,” with a range of possible values 0 to 46. For all respondents eligible for the current study, we summed the scored responses (24,25) and ranked individuals by quartiles of the aggregate score. Higher SDoH index values represented greater cumulative social disadvantage. Using quartiles of SDoH index, the first quartile group (SDoH-Q1) had the least social disadvantage and the fourth quartile group (SDoH-Q4) had the greatest social disadvantage. We performed our analyses with both the original SDoH index and the quartile categories.

**Figure 1.**
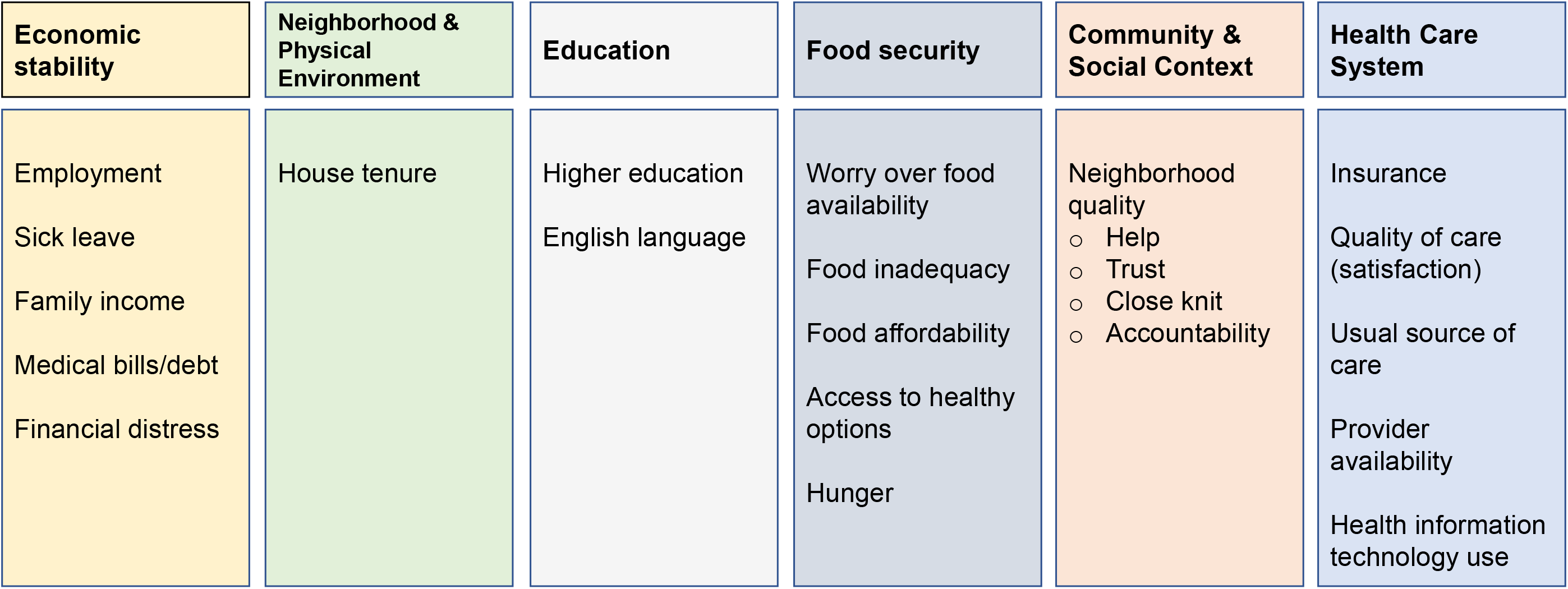
Social determinants of health from the National Health Interview Survey.

#### Dependent Variable

HALex is a validated generic HRQoL measure developed from two attributes: perceived health status and functional activity limitation. For perceived health, respondents rate their overall health as ‘poor’, ‘fair’, ‘good’, ‘very good’, or ‘excellent’. For activity limitation, each person is classified into one of six categories of activity limitation based on age and the ability to perform a major activity, as shown in **Supplemental Table S2**. Categories of physical activity limitation include “not limited in activities”, “limited in performing other activities”, “limited in performing a major activity”, “unable to perform a major activity”, “limited in instrumental activities of daily living”, and “limited in performing activities of daily living (i.e., personal care needs)” (21). Persons who are classified into more than one category are assigned to the category with the greater degree of dysfunction (21). The responses to the two items are combined in a matrix of 30 health states. The most favorable combined health state is excellent health *and* no activity limitation, and the worst state is poor health *and* an inability to perform activities of daily living. A dead state is included as the 31^st^ health state (21). Full details of the mathematical derivation of HALex – via corresponding analysis and a general multiplicative model based on multi-attribute utility theory – have been described elsewhere (21). HALex scores range from 0.10 (the worst combined health state) to 1.00 (the most favorable combined health state) for persons alive. Dead state has a value of 0 (21).

We evaluated HALex as both a continuous outcome (HALex score) and a binary outcome, poor HALex (yes/no). Poor HALex was defined as a HALex score less than the 20^th^ percentile (0.79) for the analytic sample (26). The minimal clinically important difference has not been established for HALex, but studies estimate a difference of 0.03 to be clinically relevant (27,28); we interpreted our results in light of this difference. Respondents (8.0%) with missing HALex values (i.e., with incomplete responses to at least one of the two attribute items for HALex) were excluded from the analytic sample.

#### Covariates

Covariates included age, sex, race/ethnicity, smoking status, and 11 self-reported comorbidities. Regarding comorbidities, participants reported ever receiving a clinician diagnosis of arthritis, cancer, chronic liver disease, angina or myocardial infarction (referred collectively as coronary heart disease), chronic obstructive pulmonary disease (COPD), diabetes mellitus, hypertension, and stroke. They also reported a diagnosis of kidney failure in the twelve months before the interview. Finally, respondents recalled their weights and heights from which body-mass indices were estimated, and indices ≥30 kg/m^2^ were classified as obesity.

### Statistical Analysis

We examined respondent demographics (age, sex, race/ethnicity) and health (smoking, comorbidities) characteristics across SDoH quartiles. Continuous variables were summarized as mean (SD) or median (IQR), where appropriate, and categorical variables were presented as frequencies with weighted proportions. Analysis of variance and chi-squared statistics were used to compare continuous variables and categorical variables across SDoH quartiles, respectively. We compared the distribution of HALex scores and the prevalence of poor HALex by SDoH quartiles, overall and for age, sex, and racial/ethnic categories.

For primary analyses, we evaluated both HALex outcomes first using the original SDoH index and then quartile groups of the index as independent variables. HALex scores were converted onto a decrement utility scale using negative linear transformation (X = 1 – U, where U = utility index) before assessment with two-part modeling (29). We first modelled the probability that a person had a non-zero transformed score with a logit model using the full sample (first part). In the second part, we used an ordinary least square regression model to estimate the predicted difference in the transformed scores using the subset of people with non-zero scores (29). To obtain predicted estimates and confidence limits on the original HALex scale, we simply back-transformed (1 – X) the regression estimates (29). Poor HALex was analyzed using odds ratio (OR) estimates from logistic regression. For both outcomes, three sequential models were tested: an unadjusted model; Model 1 adjusted for age (continuous), sex, and race/ethnicity; and Model 2 further adjusted for smoking and the comorbidities.

We performed two secondary analyses. First, we stratified our analysis of the outcomes by age, sex, and race/ethnicity categories. For models that utilized the continuous SDoH index as the independent variable, estimates for a 3-unit increment in the index were reported for a clearer appreciation of the differences in predicted HALex score changes across the levels of the stratifying variable. Next, we reanalyzed the cumulative SDoH-HALex association after multiply imputing by chained equations the missing values for variables with considerable (>5%) missingness – close-knit neighborhood (5.2%), helping neighbors (5.4%), trusting neighbors (5.6%), sick leave provision (6.4%), family income level (7.3%), difficulty paying medical bills (7.6%), and English language proficiency (10.2%). Independent variables for the imputation models included all covariates in the full model of the primary analyses, HALex score, and the following SDoH-related variables selected with subject-matter knowledge □ employment status, worry about food running out, educational attainment, and insurance status. Twenty complete datasets were created.

All statistical analyses incorporated the complex survey design and weighting for selection probabilities and non-response. Variance estimation for the entire pooled cohort was obtained from the Integrated Public Use Microdata Series (https://nhis.ipums.org/nhis/). Statistical significance was assessed with a two-tailed significance level of 5%. We used Stata version 16 software (StataCorp, College Station, Texas) for all analyses.

## Results

The analytic sample comprised 156,182 NHIS adult participants (mean age 46 years [SD 17]; 51.7% women) with no missing HALex scores, which represented 232.8 million U.S. adults from 2013 to 2017. Persons in higher quartile groups of the SDoH index were younger, more likely to be non-Hispanic Black or Hispanic, and they more frequently reported comorbidities (**Table 1**). **Supplemental Table S3** summarizes the SDoH variables.

**Table 1.**
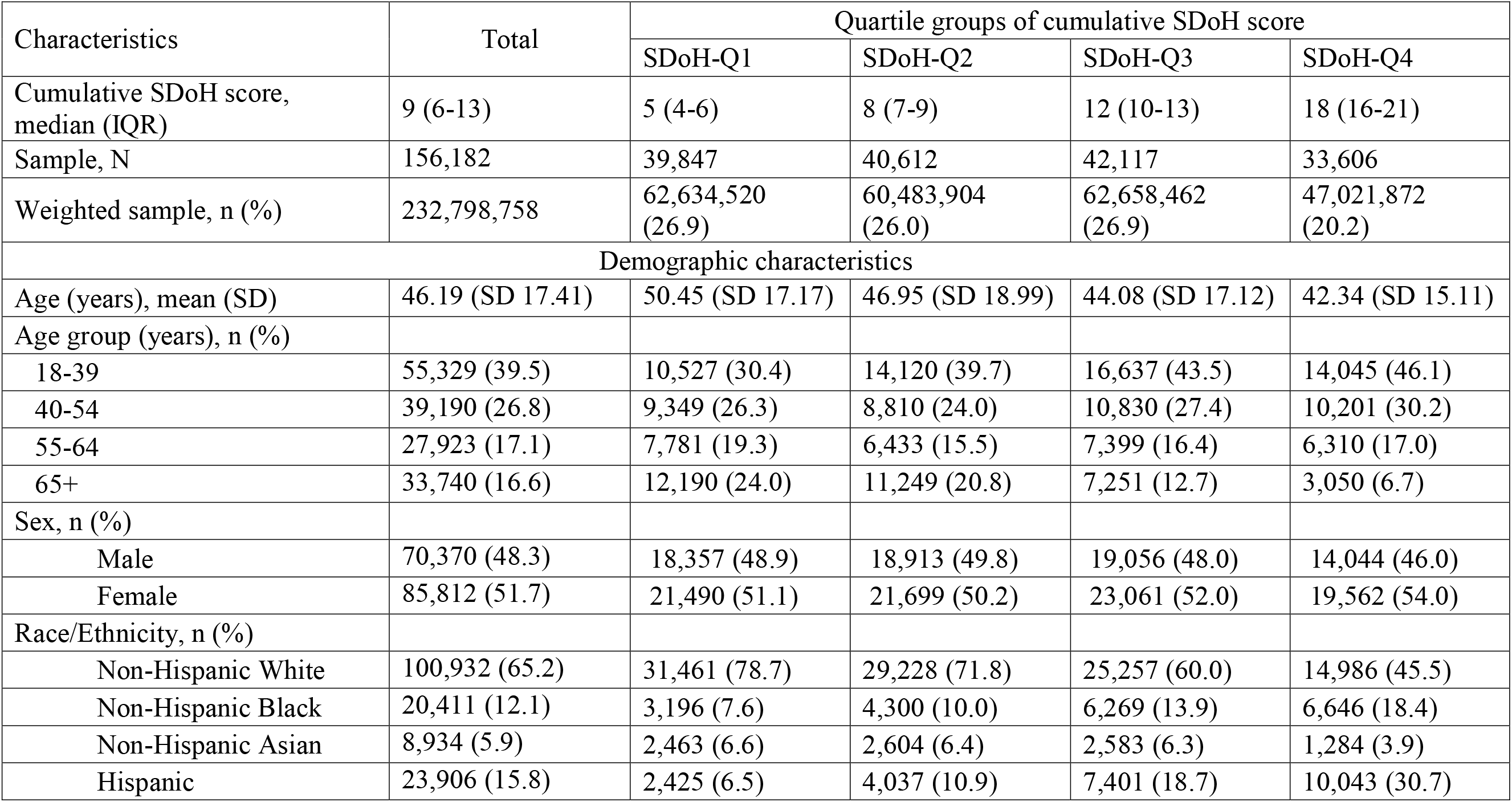
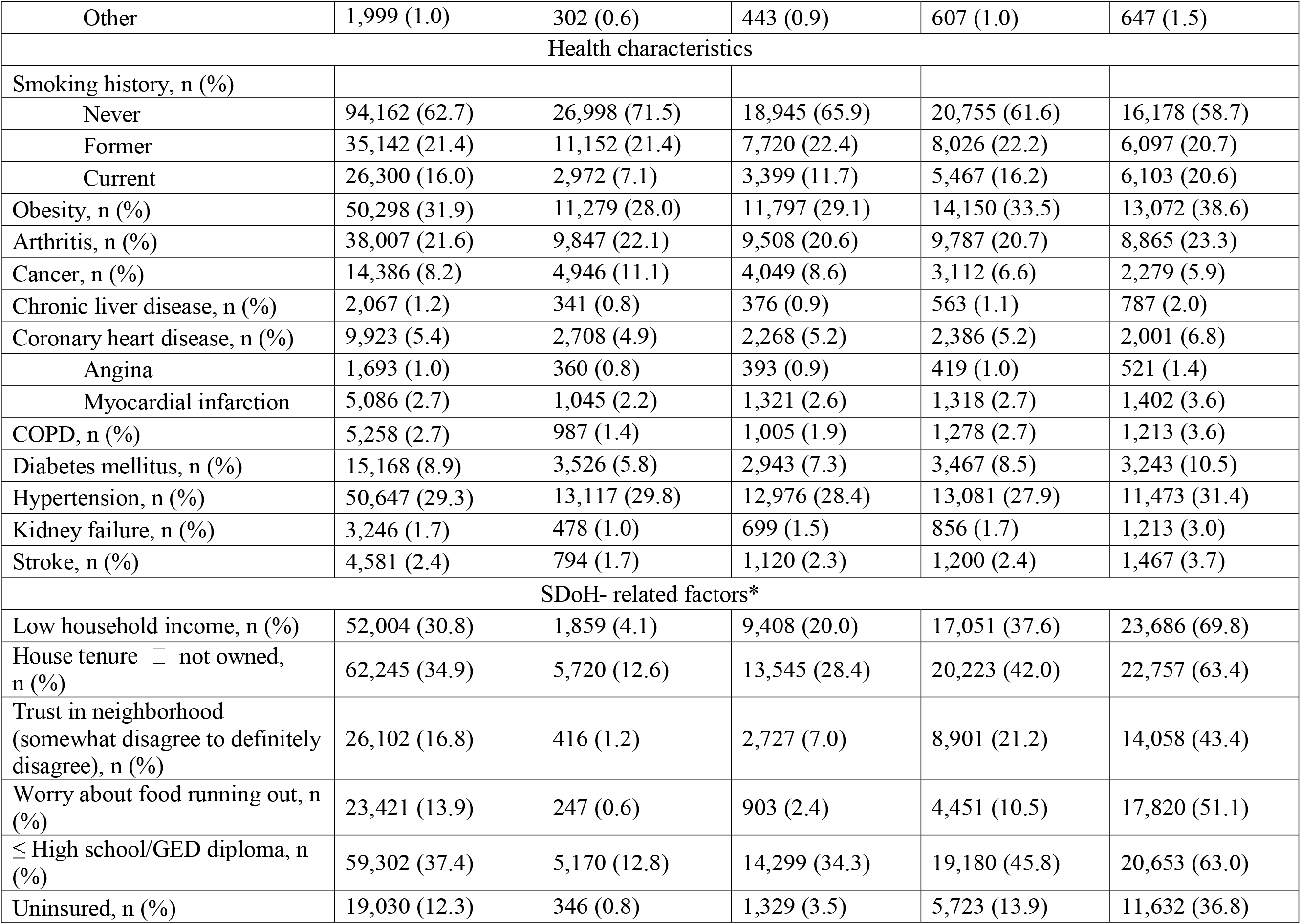
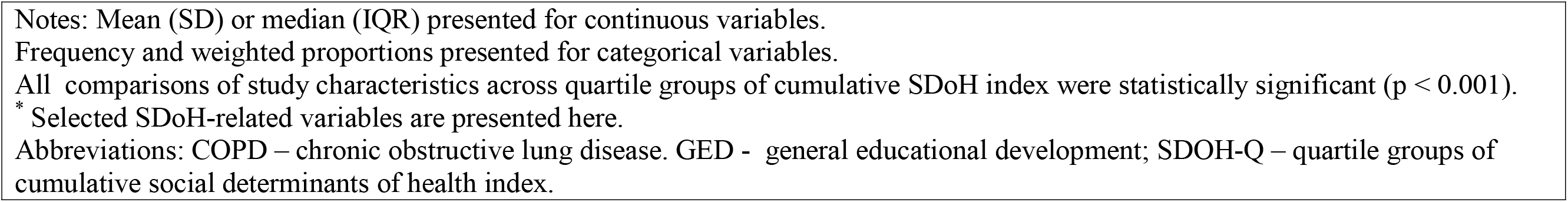
Descriptive characteristics of the adult respondents ≥18 years by quintiles of cumulative social determinants of health risk index: National Health Interview Survey 2013-2017.

| Characteristics | Total | Quartile groups of cumulative SDoH score |  |  |  |
| --- | --- | --- | --- | --- | --- |
|  |  | SDoH-Q1 | SDoH-Q2 | SDoH-Q3 | SDoH-Q4 |
| Cumulative SDoH score, median (IQR) | 9 (6-13) | 5 (4-6) | 8 (7-9) | 12 (10-13) | 18 (16-21) |
| Sample, N | 156,182 | 39,847 | 40,612 | 42,117 | 33,606 |
| Weighted sample, n (%) | 232,798,758 | 62,634,520 (26.9) | 60,483,904 (26.0) | 62,658,462 (26.9) | 47,021,872 (20.2) |
| Demographic characteristics |  |  |  |  |  |
| Age (years), mean (SD) | 46.19 (SD 17.41) | 50.45 (SD 17.17) | 46.95 (SD 18.99) | 44.08 (SD 17.12) | 42.34 (SD 15.11) |
| Age group (years), n (%) |  |  |  |  |  |
| 18-39 | 55,329 (39.5) | 10,527 (30.4) | 14,120 (39.7) | 16,637 (43.5) | 14,045 (46.1) |
| 40-54 | 39,190 (26.8) | 9,349 (26.3) | 8,810 (24.0) | 10,830 (27.4) | 10,201 (30.2) |
| 55-64 | 27,923 (17.1) | 7,781 (19.3) | 6,433 (15.5) | 7,399 (16.4) | 6,310 (17.0) |
| 65+ | 33,740 (16.6) | 12,190 (24.0) | 11,249 (20.8) | 7,251 (12.7) | 3,050 (6.7) |
| Sex, n (%) |  |  |  |  |  |
| Male | 70,370 (48.3) | 18,357 (48.9) | 18,913 (49.8) | 19,056 (48.0) | 14,044 (46.0) |
| Female | 85,812 (51.7) | 21,490 (51.1) | 21,699 (50.2) | 23,061 (52.0) | 19,562 (54.0) |
| Race/Ethnicity, n (%) |  |  |  |  |  |
| Non-Hispanic White | 100,932 (65.2) | 31,461 (78.7) | 29,228 (71.8) | 25,257 (60.0) | 14,986 (45.5) |
| Non-Hispanic Black | 20,411 (12.1) | 3,196 (7.6) | 4,300 (10.0) | 6,269 (13.9) | 6,646 (18.4) |
| Non-Hispanic Asian | 8,934 (5.9) | 2,463 (6.6) | 2,604 (6.4) | 2,583 (6.3) | 1,284 (3.9) |
| Hispanic | 23,906 (15.8) | 2,425 (6.5) | 4,037 (10.9) | 7,401 (18.7) | 10,043 (30.7) |
| Other | 1,999 (1.0) | 302 (0.6) | 443 (0.9) | 607 (1.0) | 647 (1.5) |
| Health characteristics |  |  |  |  |  |
| Smoking history, n (%) |  |  |  |  |  |
| Never | 94,162 (62.7) | 26,998 (71.5) | 18,945 (65.9) | 20,755 (61.6) | 16,178 (58.7) |
| Former | 35,142 (21.4) | 11,152 (21.4) | 7,720 (22.4) | 8,026 (22.2) | 6,097 (20.7) |
| Current | 26,300 (16.0) | 2,972 (7.1) | 3,399 (11.7) | 5,467 (16.2) | 6,103 (20.6) |
| Obesity, n (%) | 50,298 (31.9) | 11,279 (28.0) | 11,797 (29.1) | 14,150 (33.5) | 13,072 (38.6) |
| Arthritis, n (%) | 38,007 (21.6) | 9,847 (22.1) | 9,508 (20.6) | 9,787 (20.7) | 8,865 (23.3) |
| Cancer, n (%) | 14,386 (8.2) | 4,946 (11.1) | 4,049 (8.6) | 3,112 (6.6) | 2,279 (5.9) |
| Chronic liver disease, n (%) | 2,067 (1.2) | 341 (0.8) | 376 (0.9) | 563 (1.1) | 787 (2.0) |
| Coronary heart disease, n (%) | 9,923 (5.4) | 2,708 (4.9) | 2,268 (5.2) | 2,386 (5.2) | 2,001 (6.8) |
| Angina | 1,693 (1.0) | 360 (0.8) | 393 (0.9) | 419 (1.0) | 521 (1.4) |
| Myocardial infarction | 5,086 (2.7) | 1,045 (2.2) | 1,321 (2.6) | 1,318 (2.7) | 1,402 (3.6) |
| COPD, n (%) | 5,258 (2.7) | 987 (1.4) | 1,005 (1.9) | 1,278 (2.7) | 1,213 (3.6) |
| Diabetes mellitus, n (%) | 15,168 (8.9) | 3,526 (5.8) | 2,943 (7.3) | 3,467 (8.5) | 3,243 (10.5) |
| Hypertension, n (%) | 50,647 (29.3) | 13,117 (29.8) | 12,976 (28.4) | 13,081 (27.9) | 11,473 (31.4) |
| Kidney failure, n (%) | 3,246 (1.7) | 478 (1.0) | 699 (1.5) | 856 (1.7) | 1,213 (3.0) |
| Stroke, n (%) | 4,581 (2.4) | 794 (1.7) | 1,120 (2.3) | 1,200 (2.4) | 1,467 (3.7) |
| SDoH- related factors* |  |  |  |  |  |
| Low household income, n (%) | 52,004 (30.8) | 1,859 (4.1) | 9,408 (20.0) | 17,051 (37.6) | 23,686 (69.8) |
| House tenure ☐ not owned, n (%) | 62,245 (34.9) | 5,720 (12.6) | 13,545 (28.4) | 20,223 (42.0) | 22,757 (63.4) |
| Trust in neighborhood (somewhat disagree to definitely disagree), n (%) | 26,102 (16.8) | 416 (1.2) | 2,727 (7.0) | 8,901 (21.2) | 14,058 (43.4) |
| Worry about food running out, n (%) | 23,421 (13.9) | 247 (0.6) | 903 (2.4) | 4,451 (10.5) | 17,820 (51.1) |
| ≤ High school/GED diploma, n (%) | 59,302 (37.4) | 5,170 (12.8) | 14,299 (34.3) | 19,180 (45.8) | 20,653 (63.0) |
| Uninsured, n (%) | 19,030 (12.3) | 346 (0.8) | 1,329 (3.5) | 5,723 (13.9) | 11,632 (36.8) |

Notes: Mean (SD) or median (IQR) presented for continuous variables.
Frequency and weighted proportions presented for categorical variables.
All comparisons of study characteristics across quartile groups of cumulative SDoH index were statistically significant ( $p < 0.001$ ).
\* Selected SDoH-related variables are presented here.
Abbreviations: COPD – chronic obstructive lung disease. GED - general educational development; SDOH-Q – quartile groups of cumulative social determinants of health index.

### Summary of HALex Scores and Poor HALex

Overall, the total population averaged an SDoH index of 10.4 (SD 5.5), and a HALex score of 0.85 (SD 0.19), with 17% performing poorly on HALex (**Supplemental Table S4**). Men and women had similar SDoH burden and performance on HALex. Despite having the least SDoH burden, persons aged ≥ 65 years averaged low HALex scores relative to their counterparts. Compared to their counterparts, individuals of Hispanic origin averaged relatively high SDoH burden (12.3 [SD 7.2]) and low HALex scores (0.79 [SD 0.26]).

Figure 2. illustrates the unadjusted mean HALex scores across SDoH quartile groups in the total population and across age, sex, and race/ethnicity categories. Overall, higher SDoH quartile groups had lower mean HALex scores. Individuals aged ≥ 65 years had the lowest HALex scores across all SDoH quartiles, while those aged 18-39 years had the highest scores. There was no apparent difference in the profile of mean HALex scores across SDoH quartiles between males and females. While non-Hispanic Asians had the most favorable HALex profile across SDoH quartiles, Hispanics had similar HALex scores as the former at higher SDoH quartiles. On the other hand, persons of Other origins had the lowest HALex scores across SDoH quartiles.

**Figure 2.**
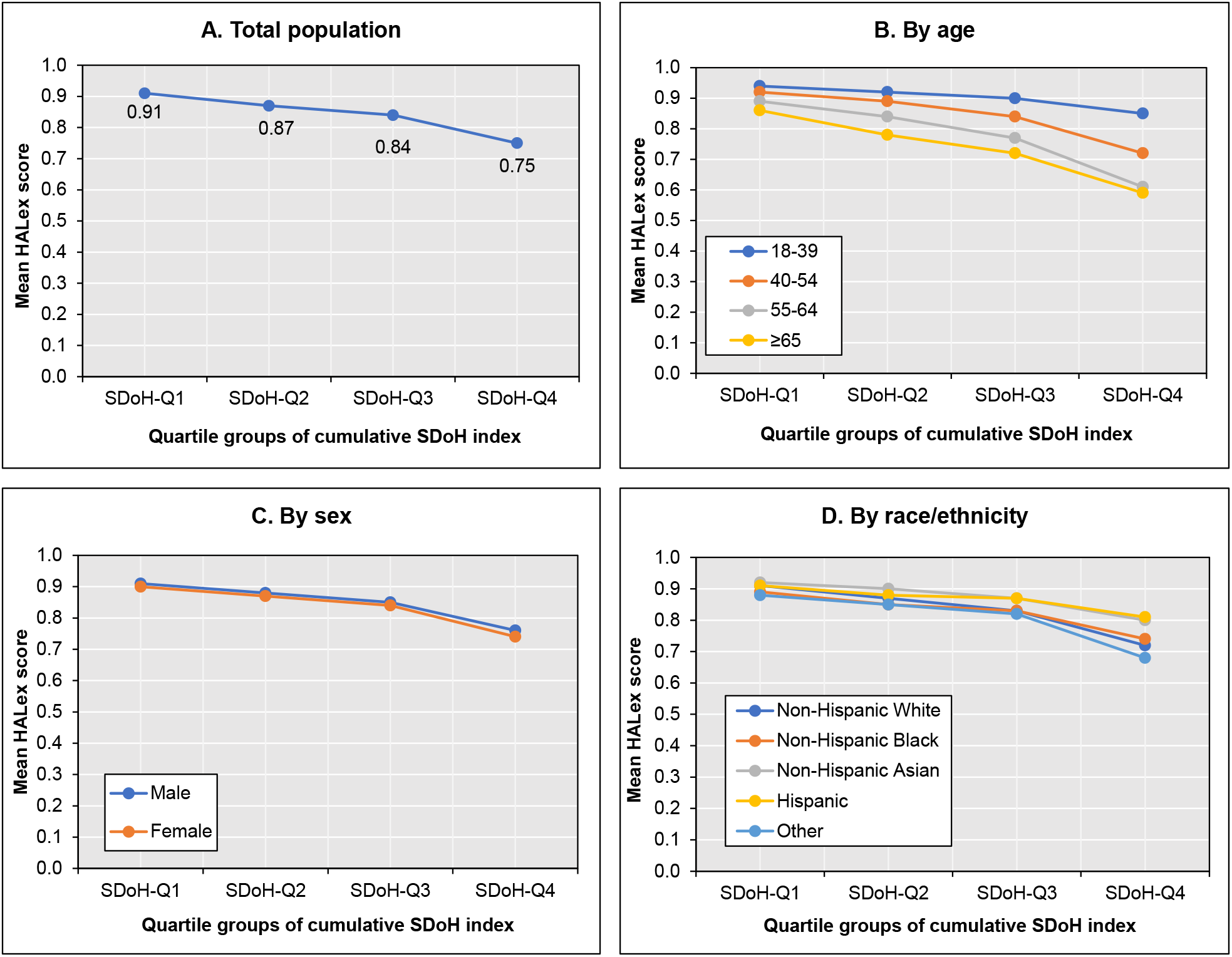
Unadjusted mean HALex scores by population characteristics. HALex – Health and Activity Limitation Index; and SDoH-Q – quartile group of cumulative social determinants of health index.

Trends in poor HALex were similar to those observed with HALex scores (**Figure 3**). The largest proportions with poor HALex were observed among persons aged ≥ 65 years, non-Hispanic White, and Other groups. Hispanic and non-Hispanic Asian groups had fewer persons with poor HALex. Males and females had similar proportions of poor HALex across SDoH quartile groups.

**Figure 3.**
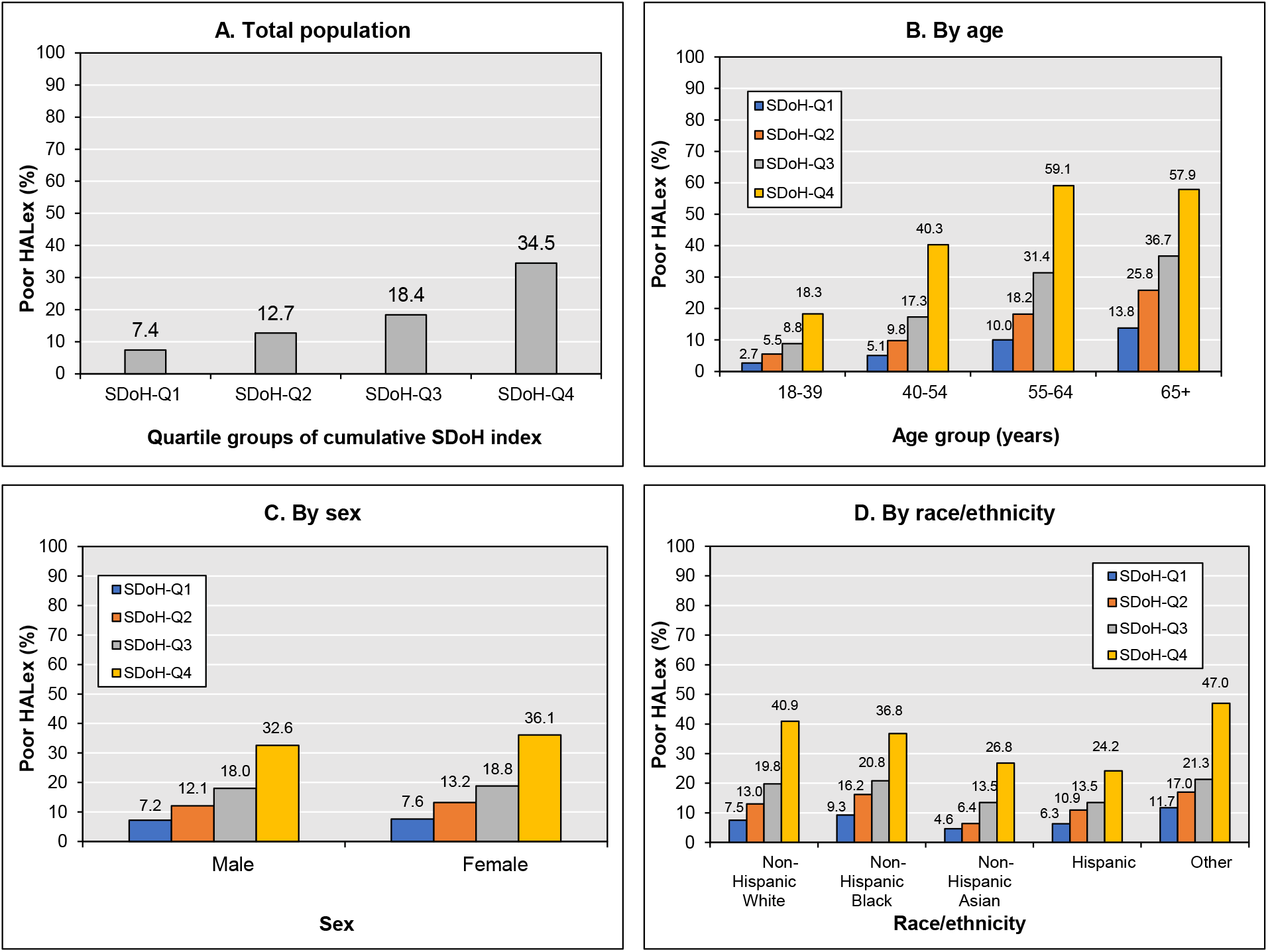
Unadjusted proportions with poor HALex by population characteristics. Poor HALex score defined by HALex score <20^th^ percentile (0.79). HALex – Health and Activity Limitation Index; and SDoH-Q – quartile group of cumulative social determinants of health index.

### Primary Multivariable Analyses

**Table 2** shows results from our multivariable regression analyses. We first report the results of the analysis of HALex scores (Table 2, Panel A). A unit increase in the SDoH index (i.e., an additional factor to cumulative social disadvantage) was associated with a decrease in HALex score (β = −0.01; 95% CI [−0.01, −0.01]) when we adjusted for age, sex, and race/ethnicity (Model 1). A similar estimate was observed after adjusting for comorbidities (Model 2). Assessing with SDoH quartiles, we observed in an incremental fashion, lower predicted HALex scores comparing higher quartile groups with SDOH-Q1, with some attenuation in the association on full adjustment. In the full model, the decrease in HALex score for SDoH-Q4 was about −0.13 (95% CI [−0.13, −0.13]).

**Table 2.**
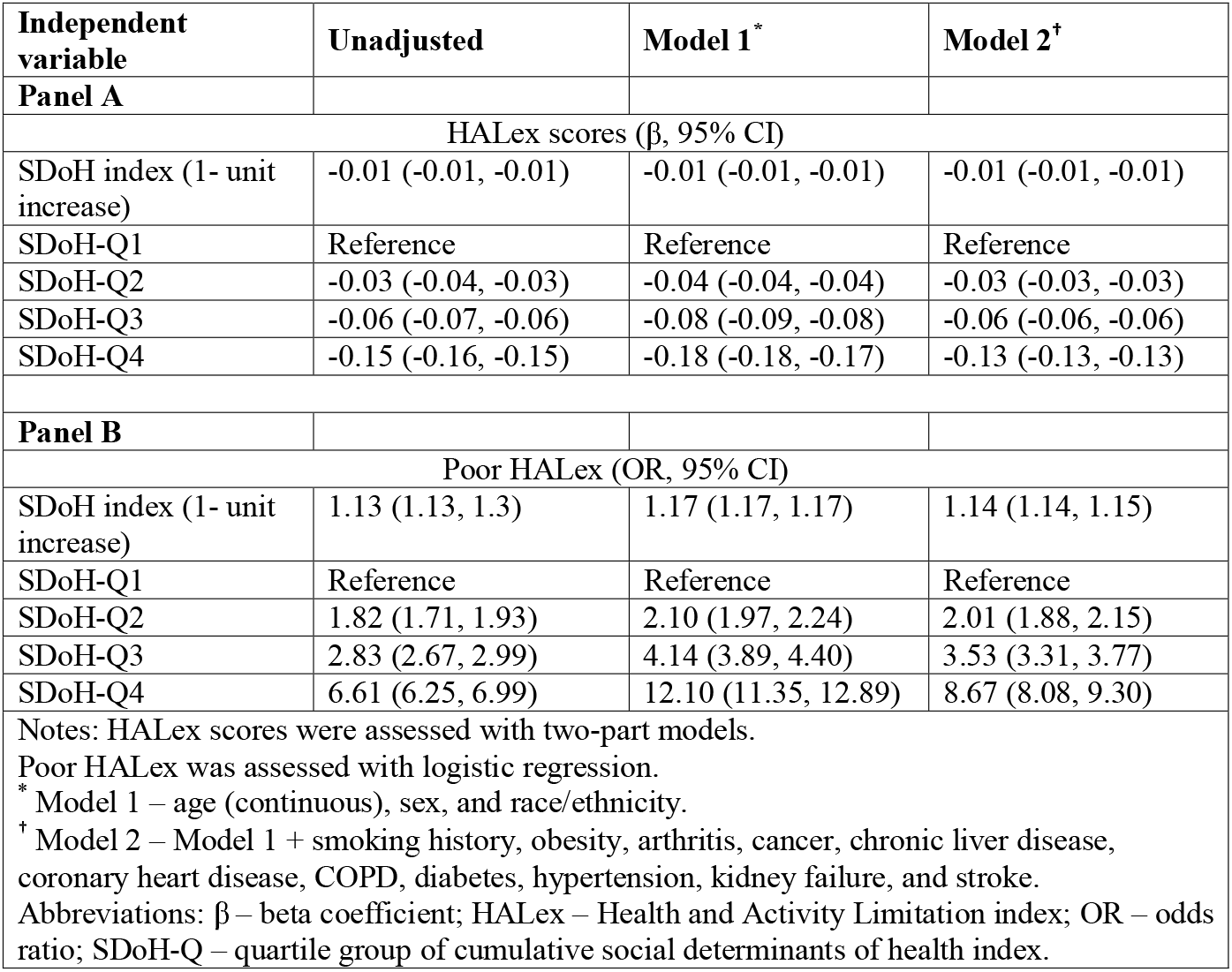
Association between cumulative social risk and the Health and Activity Limitation Index (HALex).

| Independent variable | Unadjusted | Model 1<sup>*</sup> | Model 2<sup>†</sup> |
| --- | --- | --- | --- |
| **Panel A** |  |  |  |
| HALex scores ($\beta$, 95% CI) | | | |
| SDoH index (1- unit increase) | -0.01 (-0.01, -0.01) | -0.01 (-0.01, -0.01) | -0.01 (-0.01, -0.01) |
| SDoH-Q1 | Reference | Reference | Reference |
| SDoH-Q2 | -0.03 (-0.04, -0.03) | -0.04 (-0.04, -0.04) | -0.03 (-0.03, -0.03) |
| SDoH-Q3 | -0.06 (-0.07, -0.06) | -0.08 (-0.09, -0.08) | -0.06 (-0.06, -0.06) |
| SDoH-Q4 | -0.15 (-0.16, -0.15) | -0.18 (-0.18, -0.17) | -0.13 (-0.13, -0.13) |
| **Panel B** |  |  |  |
| Poor HALex (OR, 95% CI) |  |  |  |
| SDoH index (1- unit increase) | 1.13 (1.13, 1.3) | 1.17 (1.17, 1.17) | 1.14 (1.14, 1.15) |
| SDoH-Q1 | Reference | Reference | Reference |
| SDoH-Q2 | 1.82 (1.71, 1.93) | 2.10 (1.97, 2.24) | 2.01 (1.88, 2.15) |
| SDoH-Q3 | 2.83 (2.67, 2.99) | 4.14 (3.89, 4.40) | 3.53 (3.31, 3.77) |
| SDoH-Q4 | 6.61 (6.25, 6.99) | 12.10 (11.35, 12.89) | 8.67 (8.08, 9.30) |
| Notes: HALex scores were assessed with two-part models.<br>Poor HALex was assessed with logistic regression.<br><sup>*</sup> Model 1 – age (continuous), sex, and race/ethnicity.<br><sup>†</sup> Model 2 – Model 1 + smoking history, obesity, arthritis, cancer, chronic liver disease, coronary heart disease, COPD, diabetes, hypertension, kidney failure, and stroke.<br>Abbreviations: $\beta$ – beta coefficient; HALex – Health and Activity Limitation index; OR – odds ratio; SDoH-Q – quartile group of cumulative social determinants of health index. | | | |

Regarding poor HALex (Table 2, Panel B), a unit increase in SDoH index was associated with 17% higher odds of poor HALex (OR = 1.17; 95% CI [1.17, 1.17]) in Model 1. This association barely changed when comorbidities were accounted for (OR = 1.14; 95% CI [1.14, 1.15]). Assessing with SDoH quartiles, we observed incrementally higher odds of poor HALex in both adjusted models, with some attenuation in the full model. For instance, individuals in SDoH-Q4 had 12-fold the odds of poor HALex (OR = 12.10; 95% CI [11.35, 12.89]) observed for SDoH-Q1 in Model 1. When comorbidities were accounted for, the odds of poor HALex in SDOH-Q4 were less than 9-fold that of SDoH-Q1 (OR = 8.67; 95% CI [8.08, 9.30]).

### Secondary Analyses

We first present the results of our stratified analyses for HALex scores and poor HALex, respectively (**Table 3**). The negative association between cumulative social disadvantage and HALex score was consistently observed across age, sex, and race/ethnicity subgroups. The negative cumulative SDoH-HALex association was stronger with persons aged ≥ 65 years, non-Hispanic White, and non-Hispanic Black individuals. On the other hand, Hispanic individuals had the least change in HALex scores associated with higher SDoH index. No differences in the SDoH-HALex association were observed between males and females. Trends similar to those observed with HALex score were seen in the stratified analysis of poor HALex, except the age groups. The increase in the likelihood of poor HALex with higher SDoH index or quartiles was lower among individuals aged ≥65 years than those aged 40-54 years and 55-64 years. Finally, we observed associations in the multiply imputed dataset similar to the results from our primary analyses (**Supplemental Table S4**).

**Table 3.**
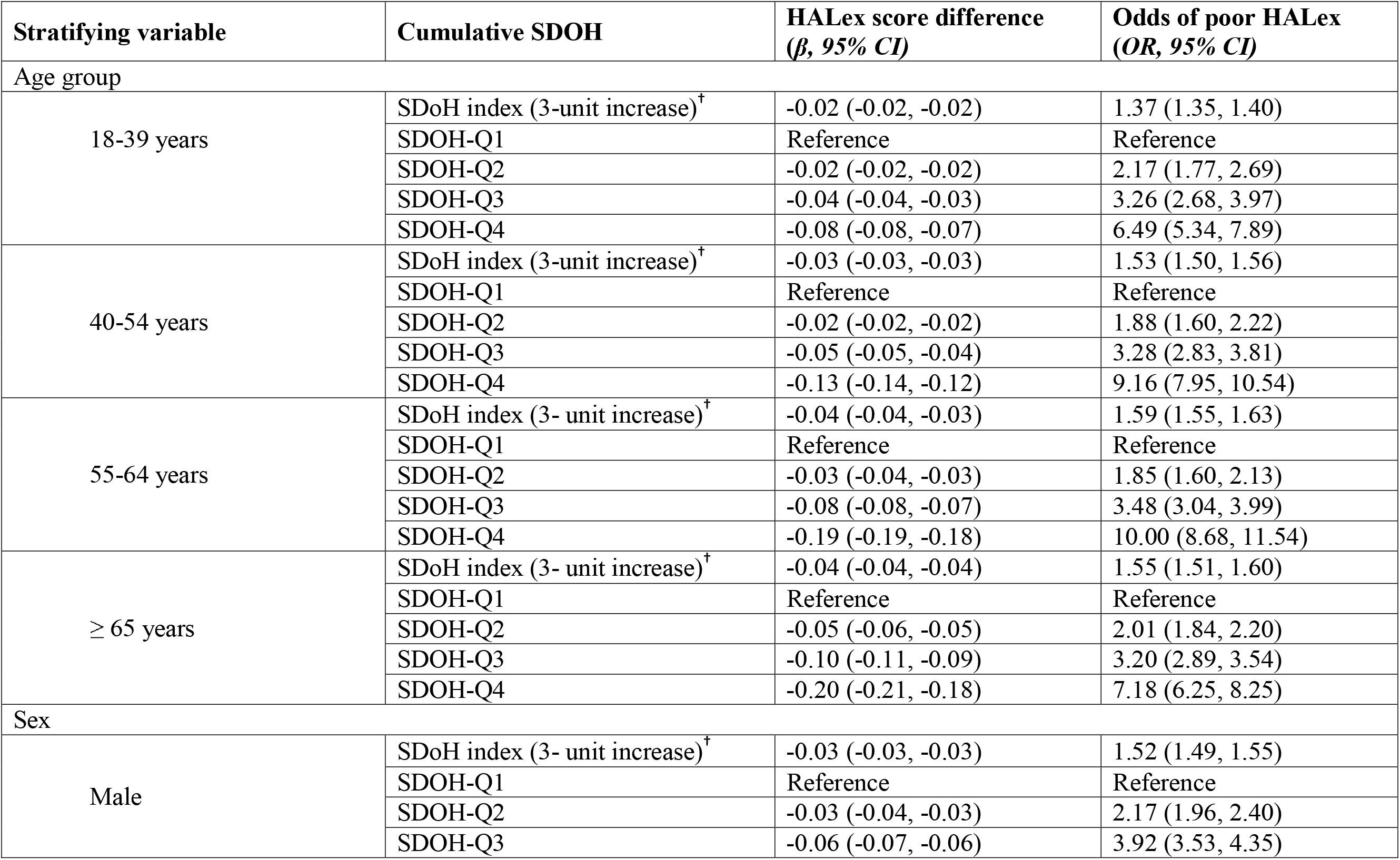
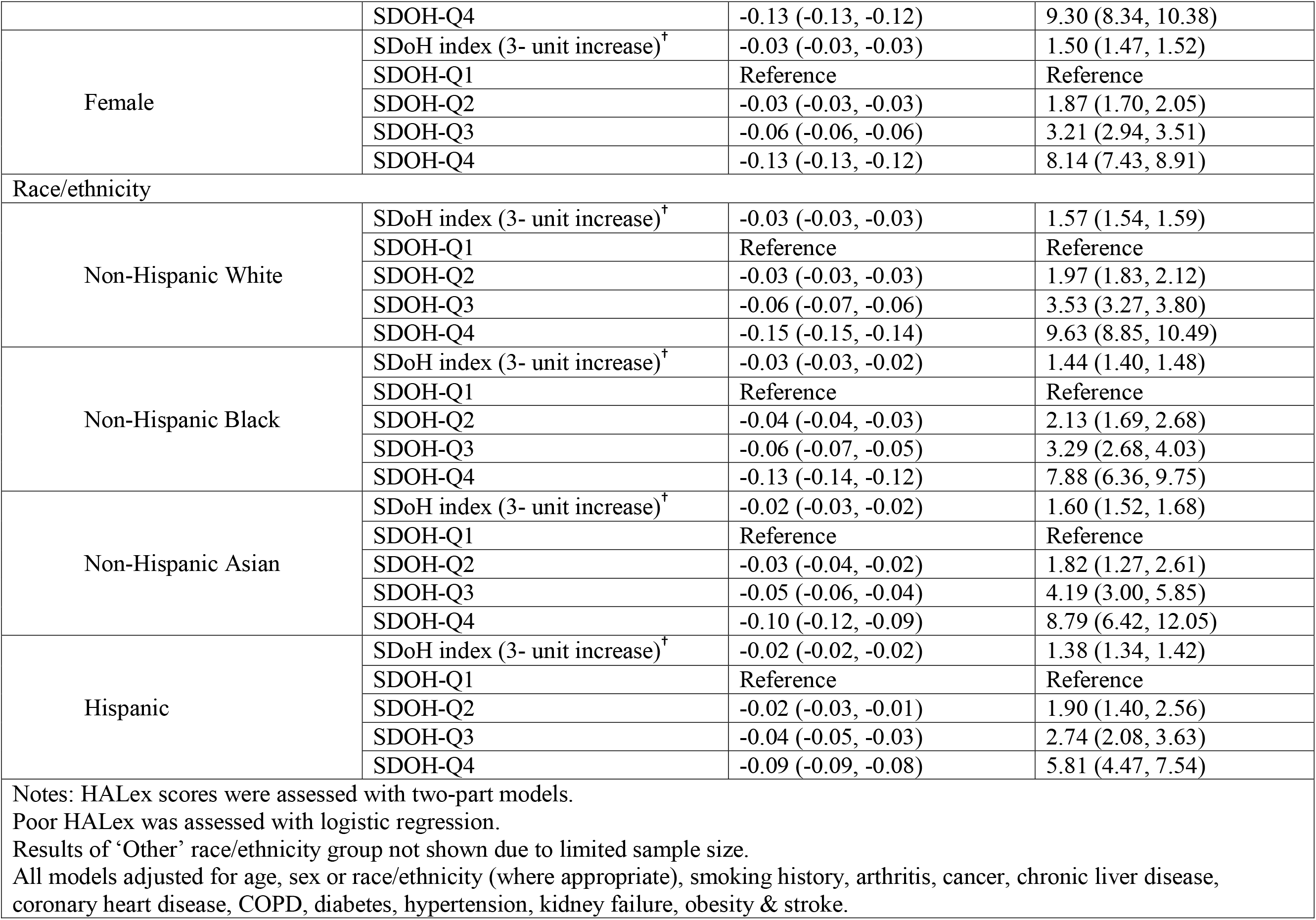
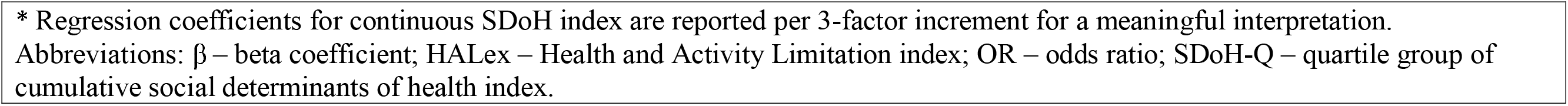
Association between cumulative SDOH risk and HALex-based outcomes, stratified by sex and race/ethnicity; National Health Interview Survey 2013-2017.

| Stratifying variable | Cumulative SDOH | HALex score difference<br>($\beta$, 95% CI) | Odds of poor HALex<br>(OR, 95% CI) |
| --- | --- | --- | --- |
| Age group |  |  |  |
| 18-39 years | SDoH index (3-unit increase)<sup>†</sup> | -0.02 (-0.02, -0.02) | 1.37 (1.35, 1.40) |
|  | SDOH-Q1 | Reference | Reference |
|  | SDOH-Q2 | -0.02 (-0.02, -0.02) | 2.17 (1.77, 2.69) |
|  | SDOH-Q3 | -0.04 (-0.04, -0.03) | 3.26 (2.68, 3.97) |
|  | SDOH-Q4 | -0.08 (-0.08, -0.07) | 6.49 (5.34, 7.89) |
| 40-54 years | SDoH index (3-unit increase)<sup>†</sup> | -0.03 (-0.03, -0.03) | 1.53 (1.50, 1.56) |
|  | SDOH-Q1 | Reference | Reference |
|  | SDOH-Q2 | -0.02 (-0.02, -0.02) | 1.88 (1.60, 2.22) |
|  | SDOH-Q3 | -0.05 (-0.05, -0.04) | 3.28 (2.83, 3.81) |
|  | SDOH-Q4 | -0.13 (-0.14, -0.12) | 9.16 (7.95, 10.54) |
| 55-64 years | SDoH index (3- unit increase)<sup>†</sup> | -0.04 (-0.04, -0.03) | 1.59 (1.55, 1.63) |
|  | SDOH-Q1 | Reference | Reference |
|  | SDOH-Q2 | -0.03 (-0.04, -0.03) | 1.85 (1.60, 2.13) |
|  | SDOH-Q3 | -0.08 (-0.08, -0.07) | 3.48 (3.04, 3.99) |
|  | SDOH-Q4 | -0.19 (-0.19, -0.18) | 10.00 (8.68, 11.54) |
| ≥ 65 years | SDoH index (3- unit increase)<sup>†</sup> | -0.04 (-0.04, -0.04) | 1.55 (1.51, 1.60) |
|  | SDOH-Q1 | Reference | Reference |
|  | SDOH-Q2 | -0.05 (-0.06, -0.05) | 2.01 (1.84, 2.20) |
|  | SDOH-Q3 | -0.10 (-0.11, -0.09) | 3.20 (2.89, 3.54) |
|  | SDOH-Q4 | -0.20 (-0.21, -0.18) | 7.18 (6.25, 8.25) |
| Sex |  |  |  |
| Male | SDoH index (3- unit increase)<sup>†</sup> | -0.03 (-0.03, -0.03) | 1.52 (1.49, 1.55) |
|  | SDOH-Q1 | Reference | Reference |
|  | SDOH-Q2 | -0.03 (-0.04, -0.03) | 2.17 (1.96, 2.40) |
|  | SDOH-Q3 | -0.06 (-0.07, -0.06) | 3.92 (3.53, 4.35) |
|  | SDOH-Q4 | -0.13 (-0.13, -0.12) | 9.30 (8.34, 10.38) |
| Female | SDoH index (3- unit increase)<sup>†</sup> | -0.03 (-0.03, -0.03) | 1.50 (1.47, 1.52) |
|  | SDOH-Q1 | Reference | Reference |
|  | SDOH-Q2 | -0.03 (-0.03, -0.03) | 1.87 (1.70, 2.05) |
|  | SDOH-Q3 | -0.06 (-0.06, -0.06) | 3.21 (2.94, 3.51) |
|  | SDOH-Q4 | -0.13 (-0.13, -0.12) | 8.14 (7.43, 8.91) |
| Race/ethnicity |  |  |  |
| Non-Hispanic White | SDoH index (3- unit increase)<sup>†</sup> | -0.03 (-0.03, -0.03) | 1.57 (1.54, 1.59) |
|  | SDOH-Q1 | Reference | Reference |
|  | SDOH-Q2 | -0.03 (-0.03, -0.03) | 1.97 (1.83, 2.12) |
|  | SDOH-Q3 | -0.06 (-0.07, -0.06) | 3.53 (3.27, 3.80) |
|  | SDOH-Q4 | -0.15 (-0.15, -0.14) | 9.63 (8.85, 10.49) |
| Non-Hispanic Black | SDoH index (3- unit increase)<sup>†</sup> | -0.03 (-0.03, -0.02) | 1.44 (1.40, 1.48) |
|  | SDOH-Q1 | Reference | Reference |
|  | SDOH-Q2 | -0.04 (-0.04, -0.03) | 2.13 (1.69, 2.68) |
|  | SDOH-Q3 | -0.06 (-0.07, -0.05) | 3.29 (2.68, 4.03) |
|  | SDOH-Q4 | -0.13 (-0.14, -0.12) | 7.88 (6.36, 9.75) |
| Non-Hispanic Asian | SDoH index (3- unit increase)<sup>†</sup> | -0.02 (-0.03, -0.02) | 1.60 (1.52, 1.68) |
|  | SDOH-Q1 | Reference | Reference |
|  | SDOH-Q2 | -0.03 (-0.04, -0.02) | 1.82 (1.27, 2.61) |
|  | SDOH-Q3 | -0.05 (-0.06, -0.04) | 4.19 (3.00, 5.85) |
|  | SDOH-Q4 | -0.10 (-0.12, -0.09) | 8.79 (6.42, 12.05) |
| Hispanic | SDoH index (3- unit increase)<sup>†</sup> | -0.02 (-0.02, -0.02) | 1.38 (1.34, 1.42) |
|  | SDOH-Q1 | Reference | Reference |
|  | SDOH-Q2 | -0.02 (-0.03, -0.01) | 1.90 (1.40, 2.56) |
|  | SDOH-Q3 | -0.04 (-0.05, -0.03) | 2.74 (2.08, 3.63) |
|  | SDOH-Q4 | -0.09 (-0.09, -0.08) | 5.81 (4.47, 7.54) |

Notes: HALex scores were assessed with two-part models.
Poor HALex was assessed with logistic regression.
Results of 'Other' race/ethnicity group not shown due to limited sample size.
All models adjusted for age, sex or race/ethnicity (where appropriate), smoking history, arthritis, cancer, chronic liver disease, coronary heart disease, COPD, diabetes, hypertension, kidney failure, obesity & stroke.
\* Regression coefficients for continuous SDoH index are reported per 3-factor increment for a meaningful interpretation. Abbreviations: $\beta$ – beta coefficient; HALex – Health and Activity Limitation index; OR – odds ratio; SDoH-Q – quartile group of cumulative social determinants of health index.

## Discussion

In this nationally representative study of U.S. adults, we demonstrated that the simultaneous accumulation of adverse socioeconomic factors across 6 domains of SDoH – economic stability, neighborhood quality, education, food security, social cohesion, and healthcare system – is incrementally associated with lower HRQoL. Accounting for demographic and clinical characteristics did not substantially attenuate this negative association.

Cumulative disadvantage may refer to two distinct processes. On one hand, it may refer to a life-course (historical and persistent) exposure to socioeconomic events and roles (30). On the other hand, and relevant to the present study, it implies the aggregation of individual socioeconomic factors in a snapshot (12). Although previous studies have demonstrated single measures of socioeconomic disadvantage at the individual and community levels to be associated with lower HRQoL (31–34), fewer had evaluated their cumulative contributions (17,35,36). Our findings expand the literature in this field by using a 47-variable construct of cumulative social disadvantage to assess its association with HALex, a generic HRQoL measure with construct validity and low administrative burden. Unlike our comprehensive assessment of social disadvantage, previous studies assessed cumulative social disadvantage with a limited number (3 to 10) of factors (17,37). Additionally, existing literature have only assessed the associations of cumulative social disadvantage with perceived health status and activity limitation separately (17,36). These two health assessments, though correlated, share a complex relationship and capture different aspects of health (38,39). Our use of HALex maximizes the correlation between these health assessment items, and the summary into a single index score offers an opportunity to track changes in HRQoL.

Although the minimal clinically important difference value has not been established for HALex, studies estimate a difference of 0.03 to be clinically relevant (40). With this, we observed in our analyses (Table 2) clinically relevant differences in HRQoL associated with increasing cumulative social disadvantage. Persons with adverse upstream SDoH disproportionately experience stressors related to access to safe housing, healthy food, transportation, limited English proficiency, among others (33,41). Stress from acculturation, neighborhood safety, and poor social support systems at the community level also influence the overall quality of life of people (42,43).

Minority racial and ethnic groups, with few exceptions, generally experience worse health outcomes compared to non-Hispanic White persons (44). In our study, we observed that Hispanic persons had mean HALex scores similar to non-Hispanic White persons across SDoH quartiles (Figure 1), and the weakest association between cumulative disadvantage and HALex (Table 3). In contrast, non-Hispanic White persons, despite a more favorable cumulative social disadvantage profile on average, had a greater decrease in HALex scores at similar cumulative disadvantage. The observation of Hispanic groups having health outcomes at least as favorable as non-Hispanic White groups is well reproduced in other population health studies (45–47). Of the several hypotheses proposed for this epidemiological paradox, a more resilient subjective construct of health influenced by cultural values and acceptance □ with concomitantly higher perceived health status than expected - could be relevant to our present findings (48).

On the other hand, it is also known that health expectations and perceived health status differ by socioeconomic status. Persons with favorable socioeconomic status report a greater negative impact of disease on their perceived health status and quality of life than those of lower socioeconomic status (46,49). For population health relevance, the favorable HRQoL observed in the Hispanic group draws attention to the concept of community resilience and the need for population “wellness” interventions to be responsive to such phenomenon.

We highlight the lower decreases in HALex scores and odds of poor HALex observed among elderly persons relative non-elderly persons (age <65 years). Similar age-related trends have been observed in studies that assessed HALex differences for chronic conditions like arthritis (50). A potential explanation is the phenomenon that vulnerable or aging populations tend to report better subjective health due to lowered expectations rather than ‘actual’ better health (51). This observation could also be due to the selective participation of healthier elderly persons in the survey and/or the lower participation of healthier persons in the younger age groups.

Our findings corroborate prior studies which used health utility indices elicited by other more common rating techniques like the standard gamble method (15,16). This adds to the desirability of HALex as a population health tool. Indeed, to our knowledge, this is the first large scale study to assess the association between cumulative social disadvantage and HALex. The potential of HALex to be a health monitoring tool lies in its constituent perceived health status item. With a more informed concept of health – one reinforced by patients assuming an active and shared care responsibility with their providers – perceived health status can become a reliable marker of satisfaction with treatment and wellbeing.

### Study Limitations

We note some limitations of this study. HALex as a generic HRQoL measure is not without limitations. First, as emotional, mental, and social functioning domains are omitted in HALex derivation there is a limitation of the discrimination ability of HALex (ceiling and floor effect), especially for populations who cluster at the highest level of health (45). Nevertheless, the simplicity in deriving HALex fosters its use in time- and data-constrained population research settings.

Second, the reliance of HALex on subjective health status raises the issue of how much HALex score differences associated with socioeconomic disadvantage are related to differential reporting behaviors and health expectations. Beyond latent health, a person’s rating of their health is influenced by the interplay between their sociocultural environment and biology (52,53). Such heterogeneity may explain away some of the differences we observed with the concomitant HALex. Regardless, the manner in which people account for the many dimensions of health when rating their overall health is relatively stable across specific populations (54), assuring that the incremental negative changes in HRQoL associated with increasing cumulative social disadvantage are less likely to be artificial.

Third, though a difference of 0.03 has been suggested as potentially a clinically relevant threshold for HALex (40), this has not been ascertained conclusively, limiting the interpretation of the magnitude and relevance of our estimates. However, HALex is strongly congruent with more widely used HRQoL measures (55,56) and our results are consistent with studies that used other HRQoL indices (16,31,49).

Finally, while the NHIS is designed to be representative of the US population, the reliance on self-reported health information implies some potential for variable misclassification. Nevertheless, previous studies have found a high correlation between the self-reported information in NHIS and the verified information found in other national datasets (57).

## Conclusion

In a nationally representative study of U.S. adults, cumulative social disadvantage was associated with lower HRQoL in an incremental fashion, independent of comorbidities. This association was consistent across demographic characteristics. Public health policies and interventions to address inequities in wellbeing may be augmented by using a cumulative disadvantage approach to capture the interplay of determinants and the resilience from cultural and community values in the target groups.

## Data Availability

All data produced in the present study are available upon reasonable request to the authors.

https://www.cdc.gov/nchs/nhis/1997-2018.htm

## LIST OF ABBREVIATIONS

HALex: Health and Activity Limitation Index
HRQoL: health-related quality of life
NHIS: National Health Interview Survey
OR: odds ratio
SDoH: social determinant of health
SDoH-Q: quartile groups of cumulative social determinant of health index

## DECLARATIONS

### Ethics Approval and Consent to Participate

Not applicable.

### Consent for Publication

Not applicable.

### Availability of data and materials

The datasets analyzed in the current study are available from the corresponding author on reasonable request.

### Authors’ Contributions

K.K.H., K.N., and Z.J. contributed to the study’s conception and design. Material preparation and data analysis were performed by K.K.H. and J.VE. The first draft of the manuscript was prepared by K.K.H. and Z.J. K.K.H. prepared all figures. All authors reviewed the manuscript. All authors read and approved the final manuscript.

## Acknowledgements

Not applicable.

