## Supplemental Table S1 for "Cumulative Social Disadvantage and Health-Related Quality of Life: National Health Interview Survey 2013-2017"

**SUPPLEMENTAL APPENDIX**

Table S1. Social Determinants of Health, by domains.

Table S2. Definitions of activity limitation based on information in the National Health Interview Survey.

Table S3. Distribution of social determinants of health by quartile groups of cumulative social determinants of health index.

Table S4. Association between cumulative social risk and the Health and Activity Limitation Index (HALex) with missing data imputation.

Figure S1. Distribution of cumulative SDoH index by SDoH quartile groups.

Figure S2. Distribution of HALex scores by SDoH quartile groups.

**Table S1.** Social Determinants of Health, by Domains

| ***Number*** | ***Short version of items*** | ***Long version of items*** | ***Survey Responses*** | ***Analytic recode*** | ***Missingness (%)*** |
| --- | --- | --- | --- | --- | --- |
| **ECONOMIC STABILITY** | | | | | |
| 1. | Employment | What was your employment status as of last week? | Working for pay at a job or business; With a job or business but not at work; Looking for work; Working, but not for pay, at a family-owned job or business; Not working at a job or business and not looking for work | 0 = "Employed or Retired"; 1 = "Never or Previously Employed" | 0.1 |
| 2. | Sick Leave | Paid sick leave at current job or most current job | Yes; No | 0 = "Yes"; 1 = "No" | 6.4 |
| 3. | Family Income | Ratio of family income to poverty threshold |  | 0 = "Middle/High-income" (≥ 200% of poverty threshold); 1 = "Low-income" (< 200% of poverty threshold) | 7.3 |
| 4. | Difficulty Paying Medical Bills | In the past 12 months did you/anyone in the family have problems paying or were unable to pay any medical bills? Include bills for doctors, dentists, hospitals, therapists, medication, equipment, nursing home or home care. | Yes; No | 0 = "No"; 1 = "Yes" | 7.6 |
| 5. | Unable to Pay Medical Bills | If previous question = Yes: Do you/Does anyone in your family currently have any medical bills that you are unable to pay at all? | Yes; No | 0 = "No"; 1 = "Yes" | 0 |
| 6. | High Financial Distress Composite Score (aggregate score from the following 6 questions);  Worried about … | | From the aggregate sum of the following 6 items, divided into quartiles:  0 if < quartile 3; 1 if ≥ quartile 3 | |  |
| 7. | … Money for retirement | How worried are you right now about not having enough money for retirement? | Very worried; Moderately worried; Not too worried; Not worried at all | 0 = "Not too worried/Not worried at all"; 1 = "Mod/Very worried" | 2.7 |
| 8. | … Medical costs of illness/accident | How worried are you right now about not being able to pay medical costs of a serious illness or accident? | Very worried; Moderately worried; Not too worried; Not worried at all | 0 = "Not too worried/Not worried at all"; 1 = "Mod/Very worried" | 2.7 |
| 9. | … Maintaining standard of living | How worried are you right now about not being able to maintain the standard of living you enjoy? | Very worried; Moderately worried; Not too worried; Not worried at all | 0 = "Not too worried/Not worried at all"; 1 = "Mod/Very worried" | 2.7 |
| 10. | … Medical costs of healthcare | How worried are you right now about not being able to pay medical costs for normal healthcare? | Very worried; Moderately worried; Not too worried; Not worried at all | 0 = "Not too worried/Not worried at all"; 1 = "Mod/Very worried" | 2.7 |
| 11. | … Paying monthly bills | How worried are you right now about not having enough to pay your normal monthly bills? | Very worried; Moderately worried; Not too worried; Not worried at all | 0 = "Not too worried/Not worried at all"; 1 = "Mod/Very worried" | 2.7 |
| 12. | … Paying rent/mortgage/housing costs | How worried are you right now about not being able to pay your rent, mortgage, or other housing costs? | Very worried; Moderately worried; Not too worried; Not worried at all | 0 = "Not too worried/Not worried at all"; 1 = "Mod/Very worried" | 2.7 |
| 13. | Delayed or foregone care due to Cost | DURING THE PAST 12 MONTHS, has medical care been delayed because of worry about the cost? (Do not include dental care) | Yes; No | 0 = "No"; 1 = "Yes" | 0 |
| 14. |  | DURING THE PAST 12 MONTHS, was there any time when (31) needed medical care, but did not get it because (31) couldn't afford it? | Yes; No | 0 = "No"; 1 = "Yes" |  |
| **NEIGHBOURHOOD AND PHYSICAL ENVIRONMENT** | | | | | |
| 15. | House Tenure | Is this house/apartment owned or being bought, rented, or occupied by some other arrangement by [you/or someone in your family]? | Owned or being bought; Rented; Other arrangement | 0 = "Own or being bought"; 1 = "Rent/Other arrangement" | 0.2 |
| **COMMUNITY AND SOCIAL CONTEXT** | | | | | |
| 16. | Neighborhood Quality (Help) | How much do you agree or disagree with the following statements about your neighborhood? Would you say… People in this neighborhood help each other out. | Definitely agree; Somewhat agree; Somewhat disagree; Definitely disagree | 0 = "Agree (Somewhat/Definitely)"; 1 = "Disagree (Somewhat/Definitely)" | 5.4 |
| 17. | Neighborhood Quality (Trust) | How much do you agree or disagree with the following statements about your neighborhood? Would you say… People in this neighborhood can be trusted. | Definitely agree; Somewhat agree; Somewhat disagree; Definitely disagree | 0 = "Agree (Somewhat/Definitely)"; 1 = "Disagree (Somewhat/Definitely)" | 5.6 |
| 18. | Neighborhood Quality (Close Knit) | How much do you agree or disagree with the following statements about your neighborhood? Would you say… This is a close-knit neighborhood. | Definitely agree; Somewhat agree; Somewhat disagree; Definitely disagree | 0 = "Agree (Somewhat/Definitely)"; 1 = "Disagree (Somewhat/Definitely)" | 5.2 |
| 19. | Neighborhood Quality (Accountability) | How much do you agree or disagree with the following statements about your neighborhood? Would you say… There are people I can count on in this neighborhood. | Definitely agree; Somewhat agree; Somewhat disagree; Definitely disagree | 0 = "Agree (Somewhat/Definitely)"; 1 = "Disagree (Somewhat/Definitely)" | 5.0 |
| **FOOD SECURITY** | | | | | |
| Food Insecurity (based on US Dept. of Agriculture Standardized Questionnaire) | | | From the aggregate sum of the following 10 items:  0 = "Food Secure" (sum ≤ 2); 1 = "Food Insecure" (sum ≥ 3) | |  |
| 20. | … Worried food would run out before got money to buy more | [fill 2: I/We] worried whether [fill 3: my/our] food would run out before [fill 4: I/we] got money to buy more. Was that often true, sometimes true, or never true for [fill 1: you/your family] in the last 30 days? | Often true; Sometimes true; Never true | 0 = "Never true"; 1 = "Sometimes true/Often true" | 0 |
| 21. | … Food did not last before had money to get more | The food that [fill 1: I/we] bought just didn't last, and [fill 1: I/we] didn't have money to get more. Was that often true, sometimes true, or never true for [fill 2: you/your family] in the last 30 days? | Often true; Sometimes true; Never true | 0 = "Never true"; 1 = "Sometimes true/Often true" | 0 |
| 22. | … Could not afford to eat balanced meals | [fill 1: I/We] couldn't afford to eat balanced meals. Was that often true, sometimes true, or never true for [fill 2: you/your family] in the last 30 days? | Often true; Sometimes true; Never true | 0 = "Never true"; 1 = "Sometimes true/Often true" | 0 |
| 23. | … Cut size or skipped meals because not enough money | In the last 30 days, did [fill 1: you/you or other adults in your family] ever cut the size of your meals or skip meals because there wasn't enough money for food? | Yes; No | 0 = "No"; 1 = "Yes" | 0 |
| 24. | … If above question = Yes: How many days in past month? | In the last 30 days, how many days did this happen? | 01-30 days (continuous response) | 0 = if < 3 days; 1 = if ≥ 3 days | 0 |
| 25. | … Eat less than felt should because not enough money | In the last 30 days, did you ever eat less than you felt you should because there wasn't enough money for food? | Yes; No | 0 = "No"; 1 = "Yes" | 0 |
| 26. | … Hungry but did not eat because not enough money | In the last 30 days, were you ever hungry but didn't eat because there wasn't enough money for food? | Yes; No | 0 = "No"; 1 = "Yes" | 0 |
| 27. | … Lose weight because not enough money for food | In the last 30 days, did you lose weight because there wasn't enough money for food? | Yes; No | 0 = "No"; 1 = "Yes" | 0 |
| 28. | … Not eat for a whole day because not enough money for food | In the last 30 days, did [fill 1: you/you or other adults in your family] ever not eat for a whole day because there wasn't enough money for food? | Yes; No | 0 = "No"; 1 = "Yes" | 0 |
| 29. | … If above question = Yes: How many days in past month? | In the last 30 days, how many days did this happen? | 01-30 days (continuous response) | 0 = if < 3 days; 1 = if ≥ 3 days | 0 |
| **EDUCATION** | | | | | |
| 30. | English Language | How well do you speak English? | Very well; Well; Not well; Not at all | 0 = "Well/Very Well"; 1 = "Not well/Not at all" | 10.2 |
| 31. | Education Attainment | What is the HIGHEST level of school completed or the highest degree received? | Never attended/kindergarten only; 1st grade; 2nd grade; 3rd grade; 4th grade; 5th grade; 6th grade; 7th grade; 8th grade; 9th grade; 10th grade; 11th grade; 12th grade; GED or equivalent; High school graduate; Some college, no degree; Associate degree: occupational, technical, or vocational program; Associate degree: academic program; Bachelor's degree; Master's degree; Professional school degree; Doctoral degree | 0 = "≥ Some college"; 1 = "≤ High School" | 0.3 |
| 32. | Health Information Technology use: Looked up health info on internet | DURING THE PAST 12 MONTHS, have you ever used computers for any of the following …Look up health information on the Internet | Yes; No | 0 = "No"; 1 = "Yes" | 1.3 |
| 33. | Health Information Technology use: Filled a prescription online | DURING THE PAST 12 MONTHS, have you ever used computers for any of the following …Fill a prescription | Yes; No | 0 = "No"; 1 = "Yes" | 1.3 |
| 34. | Health Information Technology use: Scheduled a healthcare appointment online | DURING THE PAST 12 MONTHS, have you ever used computers for any of the following …Schedule an appointment with a health care provider | Yes; No | 0 = "No"; 1 = "Yes" | 1.3 |
| 35. | Health Information Technology use: Communicated with healthcare provider online | DURING THE PAST 12 MONTHS, have you ever used computers for any of the following …Communicate with a health care provider by email | Yes; No | 0 = "No"; 1 = "Yes" | 1.3 |
| 36. | Health Information Technology use: Used internet chat rooms to learn about health topics | DURING THE PAST 12 MONTHS, have you ever used computers for any of the following …Use online chat groups to learn about health topics | Yes; No | 0 = "No"; 1 = "Yes" | 1.3 |
| **HEALTHCARE SYSTEM** | | | | | |
| 37. | Insurance Status | Multiple questions | Uninsured; Private; Medicaid; Medicare; Other | 0 = "Uninsured"; 1 = "Insured" | 1.2 |
| 38. | Usual Source of Care | Is there a place that you USUALLY go to when you are sick or need advice about your health? | Yes; There is no place; There is more than one place | 0 = "Usual source of care"; 1 = "No usual source of care" | 0.7 |
| 39. | Trouble finding a doctor/provider, past 12m | DURING THE PAST 12 MONTHS, did you have any trouble finding a general doctor or provider who would see you? | Yes; No | 0 = "No"; 1 = "Yes" | 0.8 |
| 40. | MD's office not accept you as new patient, past 12m | DURING THE PAST 12 MONTHS, were you told by a doctor’s office or clinic that they would not accept you as a new patient? | Yes; No | 0 = "No"; 1 = "Yes" | 0.8 |
| 41. | MD's office not accept your insurance, past 12m | DURING THE PAST 12 MONTHS, were you told by a doctor’s office or clinic that they did not accept your health care coverage? | Yes; No | 0 = "No"; 1 = "Yes" | 0.8 |
| 42. | Delayed Medical Care: Couldn't get through on phone | There are many reasons people delay getting medical care. Have you delayed getting care for any of the following reasons in the PAST 12 MONTHS? ..... You couldn't get through on the telephone | Yes; No | 0 = "No"; 1 = "Yes" | 0.8 |
| 43. | Delayed Medical Care: Couldn't get appt soon enough | There are many reasons people delay getting medical care. Have you delayed getting care for any of the following reasons in the PAST 12 MONTHS? ..... You couldn't get an appointment soon enough | Yes; No | 0 = "No"; 1 = "Yes" | 0.8 |
| 44. | Delayed Medical Care: Wait too long at MD's office | There are many reasons people delay getting medical care. Have you delayed getting care for any of the following reasons in the PAST 12 MONTHS? ..... Once you get there, you have to wait too long to see the doctor | Yes; No | 0 = "No"; 1 = "Yes" | 0.9 |
| 45. | Delayed Medical Care: Not open when you could go | There are many reasons people delay getting medical care. Have you delayed getting care for any of the following reasons in the PAST 12 MONTHS? ..... The clinic/doctor's office wasn't open when you could get there | Yes; No | 0 = "No"; 1 = "Yes" | 0.9 |
| 46. | Delayed Medical Care: No transportation | There are many reasons people delay getting medical care. Have you delayed getting care for any of the following reasons in the PAST 12 MONTHS? ..... You didn't have transportation | Yes; No | 0 = "No"; 1 = "Yes" | 0.9 |
| 47. | Quality of Care (Satisfaction) | In general, how satisfied are you with the healthcare are you received in the past 12 months? | Very satisfied; Somewhat satisfied; Somewhat dissatisfied; Very dissatisfied; You haven't had health care in the past 12 months | 0 = "Somewhat/Very Satisfied"; 1 = "Somewhat/Very Dissatisfied or No healthcare in past year" | 2.3 |

**Table S2.** Definitions of activity limitation based on information in the National Health Interview Survey.

| Not limited | Not limited in activities regardless of age. |
| --- | --- |
| Limited in other activities | - Limited in other activities regardless of age. - Limited in activity and 65–69 years of age but able to perform ADLs and able to perform ADLs and able to perform IADLs. |
| Limited in major activity | - 64 years of age and younger – limited in amount or kind of major activity. - 65 years and older – not applicable, major activity is considered to be ADLs and IADLs. |
| Unable to perform major activity | - 64 years of age and younger – limited in amount or kind of major activity. - 65 years and older – not applicable, major activity is considered to be ADLs and IADLs. |
| Limited in IADLs | - 18–64 years of age – unable to perform routine needs without the help of other persons and unable to perform or limited in major activity. - 65 years of age and older – unable to perform routine needs without the help of other persons. |
| Limited in ADLs | - 5–64 years of age – unable to perform personal care needs without the help of other persons and unable to perform or limited in major activity. - 65 years of age and older – unable to perform personal care needs without the help of other persons. |
| IADLs include routine needs and activities, such as everyday household chores, doing necessary business, shopping, and getting around for other purposes.  ADLs include personal care needs, such as eating, bathing, dressing or getting around the home. | |

**Table S3.** Distribution of social determinants of health by quartile groups of cumulative social determinants of health index.

| **Social determinants of health** | **SDoH-Q1** | **SDoH-Q2** | **SDoH-Q3** | **SDoH-Q4** |
| --- | --- | --- | --- | --- |
| Economic stability | | | | |
| Employment – never or previously employed | 2,527 (8.3) | 6,556 (16.2) | 10,719 (24.4) | 14,484 (41.3) |
| Sick Leave – no paid sick leave | 7,734 (21.7) | 15,940 (40.2) | 20,655 (53.0) | 21,157 (69.6) |
| Family Income – low income | 1,859 (4.6) | 9,408 (18.2) | 17,051 (37.2) | 23,686 (70.4) |
| Difficulty Paying Medical Bills | 2,805 (8.7) | 6,338 (19.5) | 9,905 (27.5) | 8,522 (35.5) |
| Unable to Pay Medical Bills | 201 (0.7) | 678 (2.1) | 2,711 (7.1) | 8,314 (25.2) |
| Worried about money for retirement | 4,998 (15.6) | 13,258 (39.3) | 24,354 (61.9) | 27,063 (81.3) |
| Worried about medical costs of illness/accident | 3,211 (9.3) | 11,085 (32.1) | 23,688 (60.3) | 27,276 (82.4) |
| Worried about maintaining standard of living | 2,062 (5.9) | 8,271 (24.4) | 20,982 (53.8) | 27,329 (82.1) |
| Worried about medical costs of normal healthcare | 585 (1.7) | 3,787 (11.3) | 15,856 (41.3) | 24,576 (74.7) |
| Worried about paying monthly bills | 329 (1.1) | 2,728 (8.7) | 14,991 (39.0) | 26,651 (79.8) |
| Worried about paying rent/mortgage/housing costs | 196 (0.7) | 1,613 (5.1) | 11,117 (29.4) | 22,860 (68.7) |
| Delayed/foregone medical care due to cost | 492 (1.2) | 1,432 (3.3) | 4,739 (9.8) | 11,662 (30.6) |
| Neighborhood & Physical Environment | | | | |
| House Tenure | 5,720 (15.3) | 13,545 (29.0) | 20,223 (41.4) | 22,757 (62.1) |
| Community & Social context | | | | |
| Neighbors help - disagree | 676 (2.0) | 3,731 (9.7) | 9,443 (23.2) | 13,023 (40.1) |
| Neighbors can be trusted - disagree | 416 (1.3) | 2,727 (7.3) | 8,901 (20.9) | 14,058 (42.6) |
| Close-knit neighborhood - disagree | 5,447 (14.9) | 11,770 (31.3) | 17,653 (43.4) | 18,636 (57.5) |
| Can count on neighbors - disagree | 726 (2.1) | 3,780 (10.3) | 9,567 (23.6) | 13,257 (40.6) |
| Kessler K6 scale - high psychological distress | 72 (0.3) | 278 (0.7) | 1,098 (2.4) | 4,260 (12.2) |
| Food | | | | |
| Worried food would run out before got money to buy more | 247 (0.6) | 903 (2.5) | 4,451 (10.4) | 17,820 (51.0) |
| Food did not last before had money to get more | 154 (0.5) | 569 (1.5) | 3,331 (7.7) | 16,001 (45.5) |
| Could not afford to eat balanced meals | 157 (0.4) | 552 (1.4) | 2,886 (6.5) | 14,369 (40.2) |
| Cut size or skipped meals because not enough money | 13 (0.0) | 55 (0.1) | 696 (1.5) | 9,588 (26.1) |
| If above question = Yes: How many days in past month? - ≥3 days | 4 (0.0) | 35 (0.1) | 478 (1.1) | 7,942 (21.4) |
| Eat less than felt should because not enough money | 13 (0.0) | 69 (0.2) | 691 (1.6) | 8,635 (24.1) |
| Hungry but did not eat because not enough money | 2 (0.0) | 18 (0.0) | 255 (0.6) | 5,396 (14.8) |
| Lose weight because not enough money for food | 2 (0.0) | 9 (0.0) | 135 (0.3) | 3,272 (8.8) |
| Not eat for a whole day because not enough money for food | 0 (0.0) | 2 (0.0) | 63 (0.1) | 2,534 (6.3) |
| Education | | | | |
| English Language – does not speak well/not at all | 120 (0.3) | 708 (2.1) | 2,396 (7.1) | 4,630 (17.8) |
| Education Attainment – less than high school/GED | 5,170 (11.6) | 14,299 (31.2) | 19,180 (45.3) | 20,653 (64.2) |
| Health Information Technology use – did not look up health information on internet in the last 12 months | 12,135 (28.3) | 22,001 (49.4) | 24,045 (55.7) | 21,423 (65.5) |
| Health Information Technology use – did not fill prescription online in the last 12 months | 31,220 (82.2) | 37,773 (92.4) | 40,012 (95.0) | 32,576 (97.0) |
| Health Information Technology use – did not schedule a healthcare appointment online in the last 12 months | 30,714 (77.6) | 37,385 (90.6) | 39,663 (93.8) | 32,429 (96.7) |
| Health Information Technology use – did not communicate with healthcare provider online in the last 12 months | 29,489 (76.1) | 37,155 (90.4) | 39,557 (93.7) | 32,380 (96.5) |
| Health Information Technology use – did not use internet chat rooms to learn about health topics in the last 12 months | 36,075 (94.7) | 39,323 (96.6) | 40,718 (96.8) | 32,503 (96.9) |
| Health Care System | | | | |
| Insurance Status - uninsured | 346 (0.9) | 1,329 (3.8) | 5,723 (13.6) | 11,632 (34.0) |
| Usual Source of Care - none | 1,213 (3.5) | 3,993 (10.4) | 7,077 (16.8) | 9,294 (27.9) |
| Trouble finding a doctor/provider, past 12m | 200 (0.5) | 574 (1.4) | 1,121 (2.6) | 2,797 (7.9) |
| MD's office not accepting you as new patient, past 12m | 204 (0.5) | 533 (1.4) | 946 (2.3) | 2,260 (6.5) |
| MD's office not accepting your insurance, past 12m | 340 (0.7) | 660 (1.7) | 1,337 (3.5) | 2,693 (7.8) |
| Delayed Medical Care: Couldn't get through on phone | 165 (0.4) | 469 (1.3) | 1,071 (2.6) | 2,067 (5.7) |
| Delayed Medical Care: Couldn't get appt soon enough | 735 (1.9) | 1,568 (4.2) | 2,760 (6.7) | 4,217 (12.0) |
| Delayed Medical Care: Wait too long at MD's office | 328 (0.9) | 891 (2.2) | 1,685 (4.1) | 3,401 (10.2) |
| Delayed Medical Care: Not open when you could go | 301 (0.9) | 669 (1.8) | 1,248 (3.0) | 2,085 (5.8) |
| Delayed Medical Care: No transportation | 49 (0.1) | 186 (0.3) | 592 (1.2) | 2,508 (6.3) |
| Quality of Care (Satisfaction) – not satisfied | 954 (2.8) | 3,964 (10.8) | 8,027 (19.8) | 11,993 (35.4) |

**Table S4**. Association between cumulative social risk and the Health and Activity Limitation Index (HALex) with missing data imputation.

| **Independent variable** | **Missing values imputed*** | |
| --- | --- | --- |
|  | **Model 1** | **Model 2** |
|  | **HALex scores (β, 95% CI)** | |
| SDoH score (1-unit increase) | -0.01 (-0.01, -0.01) | -0.01 (-0.01, -0.01) |
| SDoH-Q1 | Reference | Reference |
| SDoH-Q2 | -0.06 (-0.06, -0.05) | -0.04 (-0.04, -0.04) |
| SDoH-Q3 | -0.11 (-0.11, -0.10) | -0.08 (-0.08, -0.07) |
| SDoH-Q4 | -0.20 (-0.21, -0.20) | -0.15 (-0.15, -0.14) |
|  | **Poor HALex scores (OR, 95% CI)** | |
| SDoH score (1-unit increase) | 1.17 (1.17, 1.18) | 1.15 (1.14, 1.15) |
| SDoH-Q1 | Reference | Reference |
| SDoH-Q2 | 2.49 (2.36, 2.64) | 2.31 (2.17, 2.45) |
| SDoH-Q3 | 5.20 (4.89, 5.53) | 4.31 (4.03, 4.60) |
| SDoH-Q4 | 13.64 (12.85, 14.49) | 9.58 (8.96, 10.24) |
| * Unknowns for sick leave provision, family income level, difficulty paying medical bills, and English language proficiency were imputed for 20 datasets and combined into a single set of results.  Model 1 – age (continuous), sex, and race/ethnicity.  Model 2 – Model 1 + smoking history, arthritis, cancer, chronic liver disease, coronary heart disease, COPD, diabetes, hypertension, kidney failure, obesity, and stroke.  Abbreviations: β – beta coefficient; HALex – Health and Activity Limitation index; OR – odds ratio; SDoH-Q – quartile group of cumulative social determinants of health index. | | |

**Figure S1.** Distribution of cumulative SDoH index by SDoH quartile groups

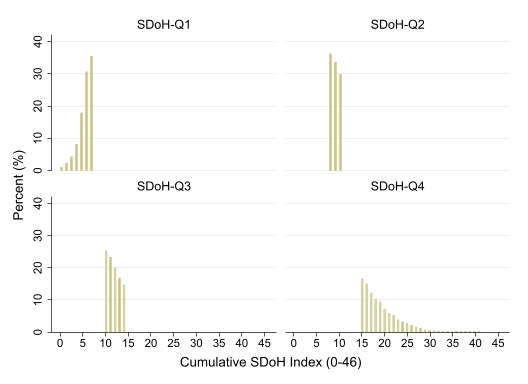

Abbreviations: SDoH-Q – quartile group of cumulative social determinants of health index.

**Figure S2.** Distribution of HALex scores by SDoH quartile groups

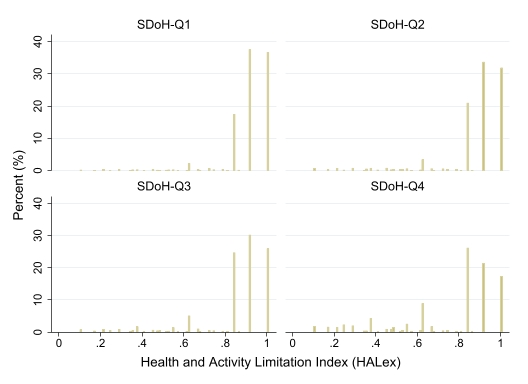

Abbreviations: SDoH-Q – quartile group of cumulative social determinants of health index.
